# AI based Chest X-Ray (CXR) Scan Texture Analysis Algorithm for Digital Test of COVID-19 Patients

**DOI:** 10.1101/2020.05.05.20091561

**Authors:** Dhurgham Al-Karawi, Shakir Al-Zaidi, Nisreen Polus, Sabah Jassim

**Affiliations:** Medical Analytica Ltd, 26A Castle Park Industrial Estate, CH6 5XA, Flint, UK; School of Computing, University of Buckingham, Buckingham, MK18 1EG, England, UK

**Keywords:** COVID-19, Chest X-Ray Scan, Artificial Intelligence, Texture Analysis, Local Binary Pattern Transform, Gabor Filter

## Abstract

Chest Imaging in COVID-19 patient management is becoming an essential tool for controlling the pandemic that is gripping the international community. It is already indicated in patients with COVID-19 and worsening respiratory status. The rapid spread of the pandemic to all continents, albeit with a nonuniform community transmission, necessitates chest imaging for medical triage of patients presenting moderate-severe clinical COVID-19 features. This paper reports the development of innovative machine learning schemes for the analysis of Chest X-Ray (CXR) scan images of COVID-19 patients in almost real-time, demonstrating significantly high accuracy in identifying COVID-19 infection. The performance testing was conducted on a combined dataset comprising CXRs of positive COVID-19 patients, patients with various viral and bacterial infections, as well as persons with a clear chest. The test resulted in successfully distinguishing CXR COVID-19 infection from the other cases with an average accuracy of **94.43%**, sensitivity **95%** and specificity **93.86%**.

**Key Strengths:** The development of efficient automatic AI texture analysis schemes for classification of chest X-Ray of COVID-19 patients with highest accuracy with equally low false negative and positive rates. Decisions would be supported by visual evidence viewable by clinician and help speed up the initial assessment process of new suspected cases, especially in a resource-constrained environment.

## 1. Introduction

The SARS-CoV-2 pandemic continues to wreak havoc across the world in a manner that unwitnessed since the unusual deadly influenza’s pandemic of 1918, otherwise known as the Spanish flu. Since December 2019 to third of May 2020, over 3.3 million cases were globally reported [1]. Despite the enormity of the present challenge, the impact of a century of gigantic advances in science, medicine and technology is manifested by the emergence of a globally adopted strategy to ensure significantly lower level of casualties. This strategy is based on (a) developing faster and more accurate diagnostic methods, (b) conducting clinical trials on possible drug treatments, and (c) developing and producing appropriate vaccines. With increasing prevalence of the disease and the nature of the outbreak, the priority is to scale up public health testing while developing new means of testing. Molecular diagnosis of COVID-19 using real-time RT-PCR test is the preferred screening/testing method [2]. While laboratory-based performance evaluations of RT-PCR test show high analytical sensitivity and near-perfect specificity with no misidentification of other coronaviruses or common respiratory pathogens, test sensitivity in clinical practice may be adversely affected by a number of variables including: adequacy of specimen, specimen type and handling, stage of infection (CDC guidelines for in-vitro diagnostics). Indeed, early reports of test performance in the Wuhan outbreak showed variable sensitivities ranging from 37% to 71% ([3], [4]).

Concerns about test availability and delays in PCR test, have led to promoting the use of serological tests for antibodies detection as proof of positive COVID-19 infection. Unlike the RT-PCR, these tests are not diagnostic testy but are meant to help estimate the proportion of the population that have had the infection and developed antibodies to it. However, recently doubts have been raised about the reliability of antibody tests. The World Health Organization’s head of emerging diseases (Dr. Maria Van Kerkhove) said that while these tests measure the level of antibodies in the blood, there is no evidence they can show “that an individual is immune or is protected against re-infection”. According to WHO, only a tiny proportion of the global population - maybe as few as 2% or 3% - appear to have antibodies in the blood showing they have been infected with COVID-19. Interested readers can find more on these issues from a variety of web reports (e.g. see [5], [6]).

Despite the many controversial views continuously reported about the reliability of antibody tests, or otherwise, they are still being developed, manufactured, and made available in some countries. For example, on 17 April 2020 - Roche (SIX: RO, ROG; OTCQX: RHHBY) announced the development and upcoming launch of its Elecsys® Anti-SARS-CoV-2 serology test to detect antibodies in people who have been exposed to the Severe Acute Respiratory Syndrome Coronavirus 2 (SARS-CoV-2) that causes the COVID-19 disease. The newly formed Rapid Testing Consortium at Oxford University have been reported to be “close to picking up 100% of all cases where people have antibodies. Now it is just a question of scaling up the manufacturing process.”

A third significant and complimentary line of investigations target the use of Artificial Intelligence based efficient classification algorithms to analyse radiology chest scan images of suspected patients. Therefore, this paper is aiming to make a contribution to these efforts.

## 2. Related Works

Radiographic chest scans with CXR and CT images, as key tools for pulmonary disease diagnosis and management, form a useful source of complementary tools for testing and management purposes in dealing with COVID-19 pandemic. Since the emergence of the disease in Wuhan, china, there has been a growing number of investigations in this direction, evidence of their use across the different diagnosis and management scenarios have been noted as of 1^st^ April 2020, in [7], as yet scant and may undergo refinement through rigorous scientific investigation. Guided by high clinical suspicion and laboratory assessment of COVID-19 suspected cases, radiographic imaging has been used to guide patient management decisions in categories of self-isolation at home, admission and isolation or evaluation for alternative diagnoses [8] and a similar process has been used in Spain [9]. Classic CXR findings in COVID-19 are multiple, bilateral, or unilateral, peripheral ground glass opacities. Organising pneumonia patterns and crazy paving may be present [10], [11].

Due to the wide availability of conventional chest radiography units especially the mobile ones and the ease of decontaminating the equipment between patients has given CXR an essential role in the fight against this pandemic. In order to support radiologist to perform such tasks, some research groups have developed Deep learning/Transfer learning techniques to analyse CXRs and classify them into COVID and non-COVID (normal CXR), however these algorithms have achieved varying results [12], [13].

On 11^th^ April 2020, M. Ilyas et al, [14], presented a review of several early AI based deep learning systems designed for the automatic detection of COVID-19 from CXR. The reported schemes are trained on different size datasets primarily to detect pneumonia as an indicator of COVID-19. With two exception all achieve considerable-to-impressive accuracy rates 80%-98%. The review concludes that all those AI approaches are detecting the subjects suffering with pneumonia without determining whether the pneumonia is caused by COVID-19 or due to any other bacterial or virus. In most cases, the training and testing datasets are rather of small size. This is a known source of overfitting by Deep learning schemes besides the black-box style of their decisions.

The main contribution of this paper is the development of innovative AI schemes for the analysis of CXR images on a combined dataset comprising CXRs of positive COVID-19 patients, patients with various viral and bacterial infections, as well as persons with clear chest. Instead of developing deep learning models, we opt for certain types of texture analysis that allow informative way of justifying the output decisions, This paper is a follow up on our recent work [15], on the analysis of Chest CT Scan Images of Coronavirus (COVID-19) Patients, and building on our accumulated experience on developing Computer Aided Diagnosis for tumour analysis we shall promote our innovative idea of texture analysis in the frequency and other transform domains.

The paper is structured as follows. Section 3 will be describing our proposed schemes, while Section 4 presents the experimental results and discussion. Section 5 concludes the existing study and illustrates some opportunities for future works.

## 3. The Proposed CXR based COVID-19 Recognition Algorithm

A computer-based system for categorising chest X-Rays scan images are expected to add innovative digital test to complement and improve performance of existing diagnostic tests for detecting COVID-19 infection in suspected patients. Integrating AI based approach into clinical practices of the RT-PCR and/or the growing number of developed antibody tests should aim to better accuracy of positive detection, hence greatly reduce the false-negative rate and improve the true positive rate. In computer vision, two different AI approaches have been dominating the research endeavour in this field, namely Deep Learning and what is loosely termed as hand-crafted feature analysis. Both share the common 2-steps strategy of extracting what is known as a “feature map” image representation followed by the use of a data classifier, and they differ only in where to put most efforts. In the DL approach, the emphasis is on extracting a feature map that encapsulate as many as possible image local features whereas the non-DL approach create a feature map consisting of one or more known/meaningful features. DL schemes require a large number of samples for training and the outcome suffers more than the non-DL schemes from overfitting [16]. In this paper, we continue to opt for non-DL approach. This is motivated by the success in our earlier work [15] on using CT scans to test for COVID-19 infection, and is justified by research done by the first and last authors, on classifying Utrasound Ovarian Tumours by AI texture analysis schemes ([17], [18], [19]). The innovative contribution of the current paper is to extend the practice of extracting image texture features from the Fourier domain into another known image transform domain, namely the Local Binary Patterns (LBP).

Our proposed AI scheme for distinguishing CXR images of COVID-19 patients from those of patients who are uninfected (i.e. normal) or infected by other viral or bacterial diseases, will be based on extracting texture features that exhibit noticeable patterns of variation relating to COVID-19 infection progression. The scheme follows the typical framework of non-deep machine learning Scheme for image classification that works by (a) pre-process the image using adaptive winner filter for noise reduction and image quality improvement, followed by inversion, (b) apply the LBP transform, (c) select a texture type and extract the appropriate feature vector, and (d) train and test the performance of each scheme on a sufficiently large dataset of image, using the linear Support Vector Machine (SVM) classifier [20].

### 3.1 Characterising Texture Features in CXR scans of COVID-19 patients

Image features refer to pixel intensity patterns or quantitative data values that reveal meaningful information about image pixels in terms of local and/or global variations. Feature extraction is the process of determining a feature vector representation of input images, and effectively isolating the most critical attributes relevant to the aim of the intended application. Popular image analysis features include colour, colour distributions, colour variation patterns from different domains (spatial or frequency domain) of an image [21]. Image-texture often refers to repeated patterns of pixel intensity value changes, and texture is accepted as a unifying approach to analysing a wide range of image types (natural, biometrics, satellite, etc.) for various tasks including segmentation, object detection/tracking, and classification.

In our classification of CT scan images of COVID-19, [15], we found that textures extracted from the Fourier transform (FFT-spectrum) provided the most COVID-19 distinguishing features. Fourier transform is a mathematical tool that represents data signals/images in terms of waveforms of different frequencies, and when applied to image data its frequency content is analysed into different ranges. Higher frequency ranges are associated with more significant texture components. The FFT spectrum quantifies the presence/strength of various frequencies.

Based on the success of the FFT-Gabor texture in achieving high accuracy for CT based analysis, we first tested the performance of this feature for our intended scheme only to find that it performed well but not as good as its performance with CT scan. Therefore, we opted to extract texture features from the LBP transform domain. The ***LBP transform*** acts as a non-linear filtered version of the image. Each pixel is replaced by a byte representing the order relation between the pixel value and its 8-immediate neighbours scanned in a clockwise manner starting from to top left corner. If the neighbour is ≥ than the centre pixel value, then the corresponding bit is set to 1 otherwise it is set to 0. See Figure 1, below for an illustration. While each pixel in the FFT spectrum depends on the entire image each in the LBP image depends on the neighbouring 8 pixels. Besides being efficient, the LBP transform is invariant to illumination (i.e. unaffected by global grey-level increase/decrease).

**Figure 1:**
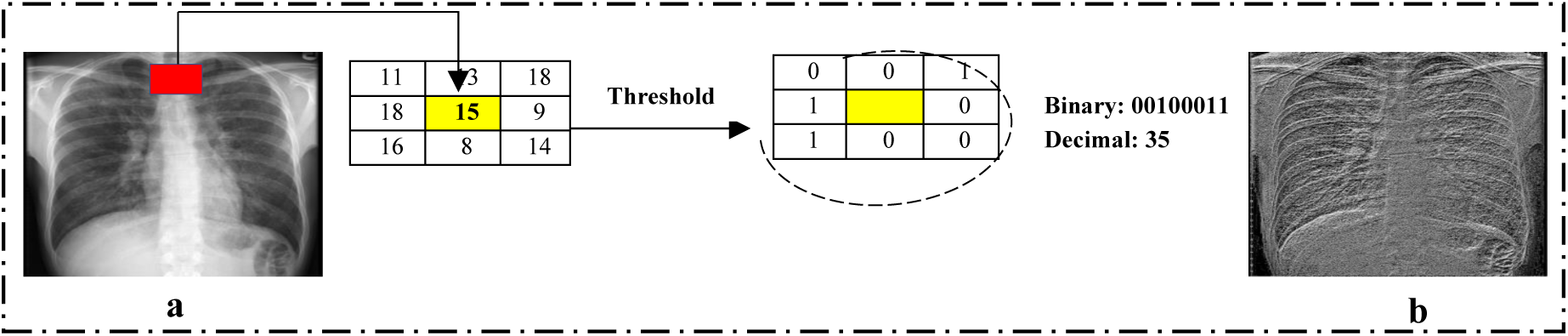
(a) Input Image with a single LBP code from 3×3 block; (b) The LBP transformed image.

We extracted various types of texture features from the LBP transformed images, but here we shall only briefly describe the best three COVID-19 discriminating texture features.

- **Gabor Texture feature**. Images are transformed by a set of parametrised 2-dimensional band-pass Gabor wavelet filters designed for detecting specific frequency information within an image region at a user chosen set of scales and orientation ranges [22]. The extracted Gabor-texture quantifies the frequency content in the input image within the sub-regions determined by the selected scales and directions.
- **The uniform LBP (59 bins)**. In many image analysis and pattern recognition applications, the 256-bins histogram of the LBP transformed images are used directly as a texture feature vector of the input images. Among the LBP codes, the uniform LBP (ULBP) codes (8-bit binary strings that contain single run of 1’s, considered as a circular string) represent pixels with significant geometric textures. There are 58 ULBP’s, and another way to form a texture feature vector representation of an input image, is a 59-bins histogram whereby where the first bin is for the non-uniform LBP’s and the remaining ones are for the 58 ULBPs [22].
- **Histograms of Oriented Gradient (HOG):** A localised texture feature, introduced in [23], that uses image oriented gradient information for detecting image objects. The oriented gradient of an image ***I*** at pixel point ***p(x,y)***, is the resultant of the horizontal vector ***g_x_ =(I(p(x+1,y))-I(p(x-1,y)))*** and the vertical vector ***g_y_ =(I(p(x,y+1))-I(p(x,y-1)))***. The image ***I*** is then subdivided into non-overlapping rectangular cells of equal size. The oriented gradient of each cell is then organised into two matrices of the same cell size, one for orientation **o *= arctan(g_y_/ g_x_)*** and the other for the magnitudes 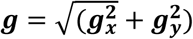. Each cell is then represented by a “***9***-bins histogram”. The bins indices represent the ***9*** orientation values ***{0, 20, 40, 60,…,160}***. For each pixel of a cell, locate the orientation of its gradient between ***2*** successive bins and split the magnitude to be added to these ***2*** bins proportionately. Finally, HOG is formed by concatenating the histograms of oriented gradients in these cells. The histograms are indicators of the location of intensity gradients and the edge directions in the image. The feature vector provides information about the shape and presence of the structures/objects within the image. In this study, images are subdivided into ***16*** rectangular cells, and HOG feature vectors size ***144=9=16***.

### 3.2 Performance Evaluation Framework of our LBP-based Schemes

This framework emulates a typical CAD process. It involves 2 key stages: training and testing. In the training stage, the feature vectors extracted from a sufficient number of samples will be used to determine the best separation of the extracted feature vectors by the chosen classifier model. Due to relatively high dimensionality of the various extracted texture feature vectors compared to the modest number of samples available for training, we choose the Linear SVM binary classifier. The output from the SVM training stage is a hyperplane that separates the feature vectors of the positive class from those of the negative class. In the testing stage, the feature vector of each testing image is extracted as usual and its signed distance from the SVM hyperplane is calculated, where predicted class is determined by the sign and actual distance is used to determine probability of the predicted class being accurate. Figure 2, below, illustrates the full process of feature extraction applied to all image at the training as well as testing stages.

**Figure 2:**
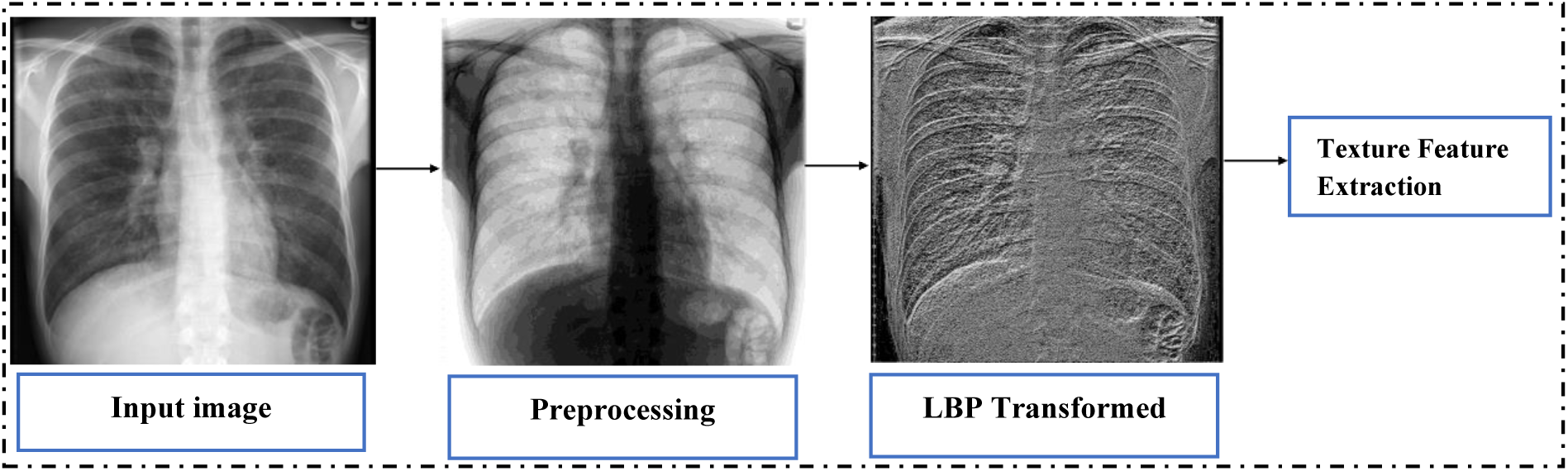
Texture Feature Extraction in the LBP Domain.

In this study, we shall conduct three different experiments: - First, COVID-19 vs. Normal, Second, COVID-19 vs. Virus Infection & Bacteria and last experiment COVID-19 vs. (Normal, Virus Infection & Bacteria all together). To test the performance of the various LBP-based schemes, for the various combinations, we assembled an experimental dataset of 1015 Chest X-ray images obtained from 2 different sources: 305 images for (COVID-19) were collected from [24], and 710 Images (Normal (305) +Viral Infection & Bacteria (405)) were collected from [25]. Figure 3, below, displays samples from these different sets.

**Figure 3:**
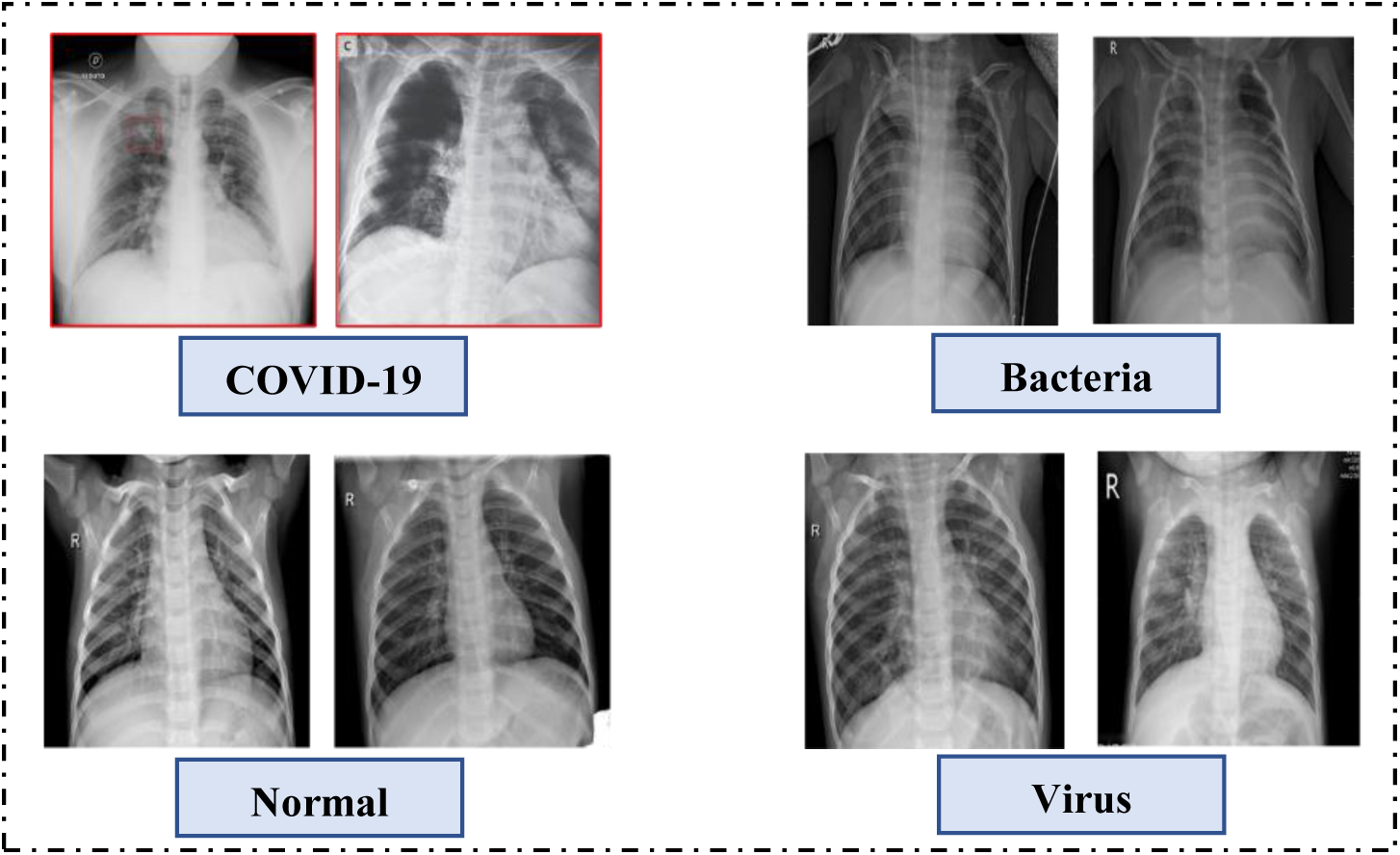
Samples of CXR images in the experimental dataset.

## 4. Experimental Protocols, Classification Results and Discussion

While the COVID-19 transition is continuing, datasets of CXR images of newly examined COVID-19 patients will be frequently updated. These updates provide us with opportunities to update, improve and hopefully soon help achieve the best performance of our schemes independent of experimental datasets, and our experimental protocols are designed to be updatable as more data sample batches become available. Accordingly, we create a dataset of “prospective test” by isolating a set of 105 of COVID-19 images and for each of the 3 different experiments, we set aside 105 from the corresponding negative class as follows:

✓ First Experiment 105 Normal CXR Images.
✓ Second Experiment 105 Virus Infection & Bacteria Images.
✓ Third Experiment 105 (35 Normal, 35 Virus Infection & 35 Bacteria) Images.

For each experiment, we shall first train and test the performance of our LBP-based schemes with the remaining images of the corresponding dataset and follow the protocol described by the left side part of the self-explanatory block diagram of Figure 4, below. At the end of this experiments, 30 SVM classifier models are output with its performance and the average of their accuracy/sensitivity/specificity rates are declared as the performance of the tested scheme. Subsequently to test any new case, we use one of the 30 SVM hyperplanes namely the model with highest accuracy rate among those for which the different between the sensitivity and specificity are minimal. This model will be used to test the performance of the scheme over the corresponding prospective testing data set, per the right-side diagram of Figure 4.

**Figure 4:**
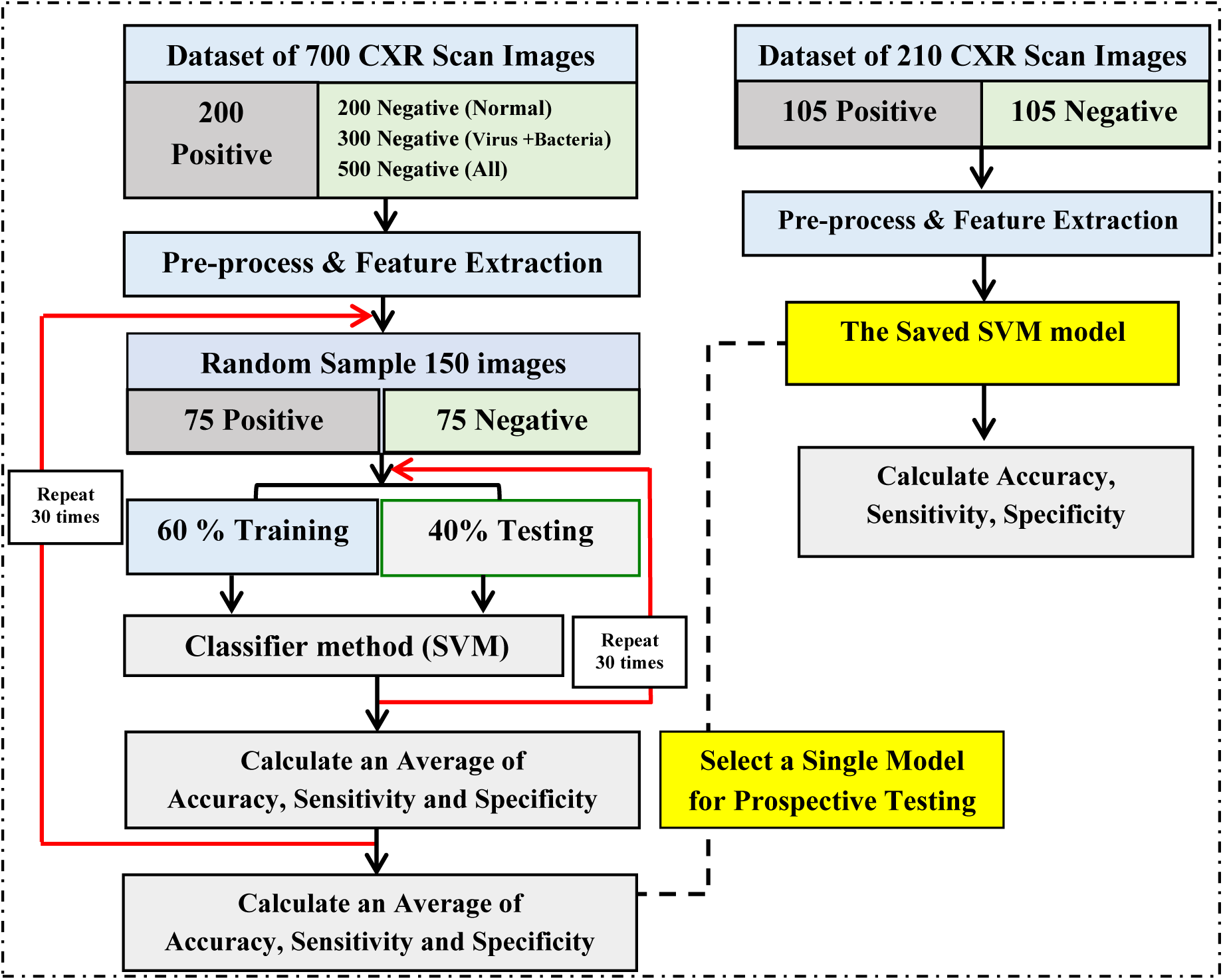
Experimental protocols for both tests.

Table 1, below, displays the results of the 3 experiments and include the performance of the 3 different LBP-based texture COVID-19 classification schemes in terms of the average accuracy, sensitivity and specificity rates for the original training/testing experiments over the 700 images dataset. In each scenario, we also present the results of fusing the three texture schemes at the decision level using simple majority rule.

**Table 1.** Classification results using SVM classifier.

| Texture Features. | Accuracy<br>($\mu$) | Sensitivity<br>($\mu$) | Specificity<br>($\mu$) |
| --- | --- | --- | --- |
| **COVID-19 VS. Normal** |  |  |  |
| **LBP-Gabor.** | **97.16%** | **96.90%** | **97.43%** |
| **LBP-HOG.** | **92.83%** | **93.52%** | **92.15%** |
| **LBP-Uniform (59 bins).** | **94.39%** | **95.29%** | **93.49%** |
| **Feature Fusion.** | **97.72%** | **97.89%** | **97.56%** |
| **COVID-19 VS. Viral Infection &Bacteria** |  |  |  |
| **LBP-Gabor.** | **92.70%** | **93.36%** | **92.05%** |
| **LBP-HOG.** | **89.94%** | **90.20%** | **89.68%** |
| **LBP-Uniform (59 bins).** | **90.72%** | **91.36%** | **90.08%** |
| **Feature Fusion.** | **92.77%** | **92.96%** | **92.59%** |
| **COVID-19 VS. Normal, Viral Infection &Bacteria all together** |  |  |  |
| **LBP-Gabor.** | **94.15%** | **95.28 %** | **93.02 %** |
| **LBP-HOG.** | **90.87 %** | **91.03 %** | **90.71 %** |
| **LBP-Uniform (59 bins).** | **91.14 %** | **90.74 %** | **91.55 %** |
| **Feature Fusion.** | **94.43%** | **95%** | **93.86 %** |

In Table 2 below, we present the performance of the saved models over the corresponding 210 prospective datasets for the 3 scenarios including the majority rule fusion results.

**Table 2.** Classification results using SVM classifier for Prospective test.

| Texture Features. | Accuracy<br>($\mu$) | Sensitivity<br>($\mu$) | Specificity<br>($\mu$) |
| --- | --- | --- | --- |
| **COVID-19 VS. Normal (Prospective Test)** |  |  |  |
| **LBP-Gabor.** | **94.69%** | **95.24%** | **94.14%** |
| **LBP-HOG.** | **90.90%** | **91.86%** | **89.95%** |
| **LBP-Uniform (59 bins).** | **92.88%** | **92.76%** | **91.01%** |
| **Feature Fusion.** | **95%** | **95.03%** | **94.98%** |
| **COVID-19 VS. Viral Infection &Bacteria (Prospective Test)** |  |  |  |
| **LBP-Gabor.** | **90.92%** | **92%** | **89.85%** |
| **LBP-HOG.** | **87.7%** | **88.28%** | **86.12%** |
| **LBP-Uniform (59 bins).** | **88.55%** | **89.72%** | **87.39%** |
| **Feature Fusion.** | **91.73%** | **92.85%** | **90.61%** |
| **COVID-19 VS. Normal, Virus Infection &Bacteria all together (Prospective Test)** |  |  |  |
| **LBP-Gabor.** | **91.76%** | **92.36 %** | **91.17 %** |
| **LBP-HOG.** | **86.97%** | **88%** | **85.96%** |
| **LBP-Uniform (59 bins).** | **90.18%** | **89.85%** | **90.52%** |
| **Feature Fusion.** | **91.95%** | **92.93%** | **91.01%** |

Overall, in each of the scenarios Table1 results show that each of the texture features has a significant power of discriminating images of COVID-19 from Normal, Virus Infection & Bacteria and all together individually. In each scenario, the LBP-Gabor scheme outperforms the other two texture schemes with the LBP-HOG being the lowest performing scheme but not by a significant amount. Perhaps, these results reinforce the widely accepted assertion that Gabor filters provide a credible mathematical model of the Human Vision system.

Furthermore, each of the 3 texture schemes achieves the highest discrimination between CXR of COVID-19 patients against normal CXR images, and the lowest discrimination is achieved between COVID-19 against all others. Finally, although fusion of the 3 LBP-texture schemes resulted in improved performance, unfortunately only marginally. This may be explained as a shortcoming of the primitive majority decision rule rather than fusion in general, but it also reflects the superiority of the LBP-Gabor scheme which distinguishes COVID-19 CXR’s from the normal, viral bacterial, and All cases with accuracy of **97.16%**, **92.70%**, and **94.15%** respectively. In all scenarios, the sensitivity and specificity are close to the accuracy rates. It is worth noting that extracting Gabor features in the CXR spatial domain achieved significantly lower accuracy for the same experimental data set.

These results reveal that the Gabor filtering in the LBP transform uncovers all significantly visible texture features in infected CXRs throughout the chest that are not limited to the Ribs or the Chest cage bones. The additional visible features are reflected as artefacts that must have been caused by X-Rays detectible infection-caused changes in the chest and airways. These observations are clearly illustrated in the appendix displayed images, especially in the off-diagonal top-left Gabor sub-images. These artefacts are not hardly visible in the normal CXRs. Moreover, we observe notable differences in the extent of additional artefacts between the CXR’s of COVID-19 infected patients from those of the patients exhibiting viral and Bacterial infection. These observations, if further confirmed, maybe useful in providing the clinician with convincing evidences of the LBP-Gabor scheme predicted decision by complementing our interface software with a visual display of some of the created sub-images.

The results in Table 2 for the prospective tests follow the same pattern described above, but in all cases, there is a tolerable reduction in accuracy for all scenarios. This reduction can be explained by the fact none of the images in the prospective datasets is involved in the training/testing of the SVM model. The fact that all features were extracted directly from the LBP images with image enhancement and de-noising, demonstrate the success of our innovative strategy of texture analysis in the LBP transform domain. It is worth noting that, we test a larger number of LBP-texture schemes and most of the unpresented schemes achieved in-excess of 80% accuracy.

## 5. Conclusion and Feature Works

The paper investigated and evaluated our new innovative approach of extracting texture features from the LBP-transformed CXR images of different types of Chest infections, with the aim of developing an automatic CAD system to be used for distinguishing between COVID-19 and Non COVID-19. This paper is a follow up on our COVID-19 infection detection by texture analysis CT scan images where the FFT-Gabor scheme achieved accuracy in-excess of **95%**. The choice of using CXR instead of CT (standard and Low dose) is due to the significant differences in the harmful ionising radiation dose (0.1 mSv for CXR compared to 7 mSv for standard CT and 1.5 mSv for LDCT). Consequently, CXR is a much safer option. Experimental test results over different collected sufficiently large datasets show that the corresponding SVM models built on several texture features extracted from the LBP domain achieved accuracy well above random guess (min 75%). The LBP-Gabor filters attained the highest and stable accuracy of **94.15%** with equally impressive near equal sensitivity and specificity rates. Decision level fusion of all texture features resulted in average accuracies close to **95%**, and higher than those by the individual features. These results demonstrate the viability and effectiveness of the LBP transform domain to extracted features for the stated task. We have also observed the possibility of using the LBP-Gabor filtered sub-images as an additional visual evidence of the predicted decision within clinical setting. Due to the promising results, we are currently testing LBP-Gabor and FFT-Gabor texture features for Lung tumour detection.

## Data Availability

public Datasets

https://github.com/ieee8023/covid-chestxray-dataset/tree/master/images,

https://data.mendeley.com/datasets/rscbjbr9sj/3

## Appendix

Below is an illustration of the output of using the LBP-Gabor scheme that filters the LBP transformed images (2 Normal and 2 COVID-19) in 5 scales and 10 orientations. It is easy to visibly note the differences in the pattern displayed in each of the 50 small LBP-Gabor blocks between the COVID-19 and normal cases. Such displays provide the clinician and informative justification of the output decision.

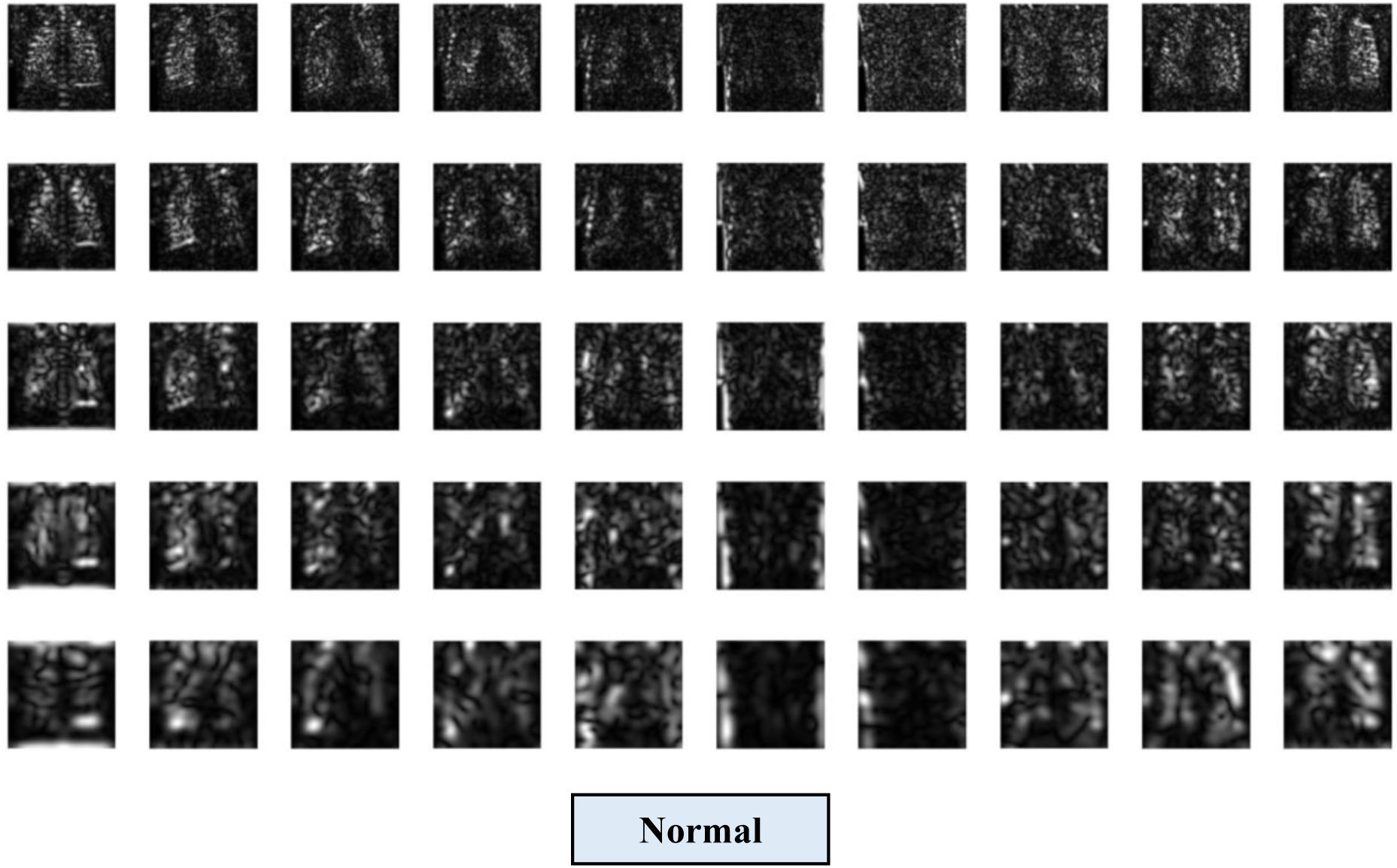

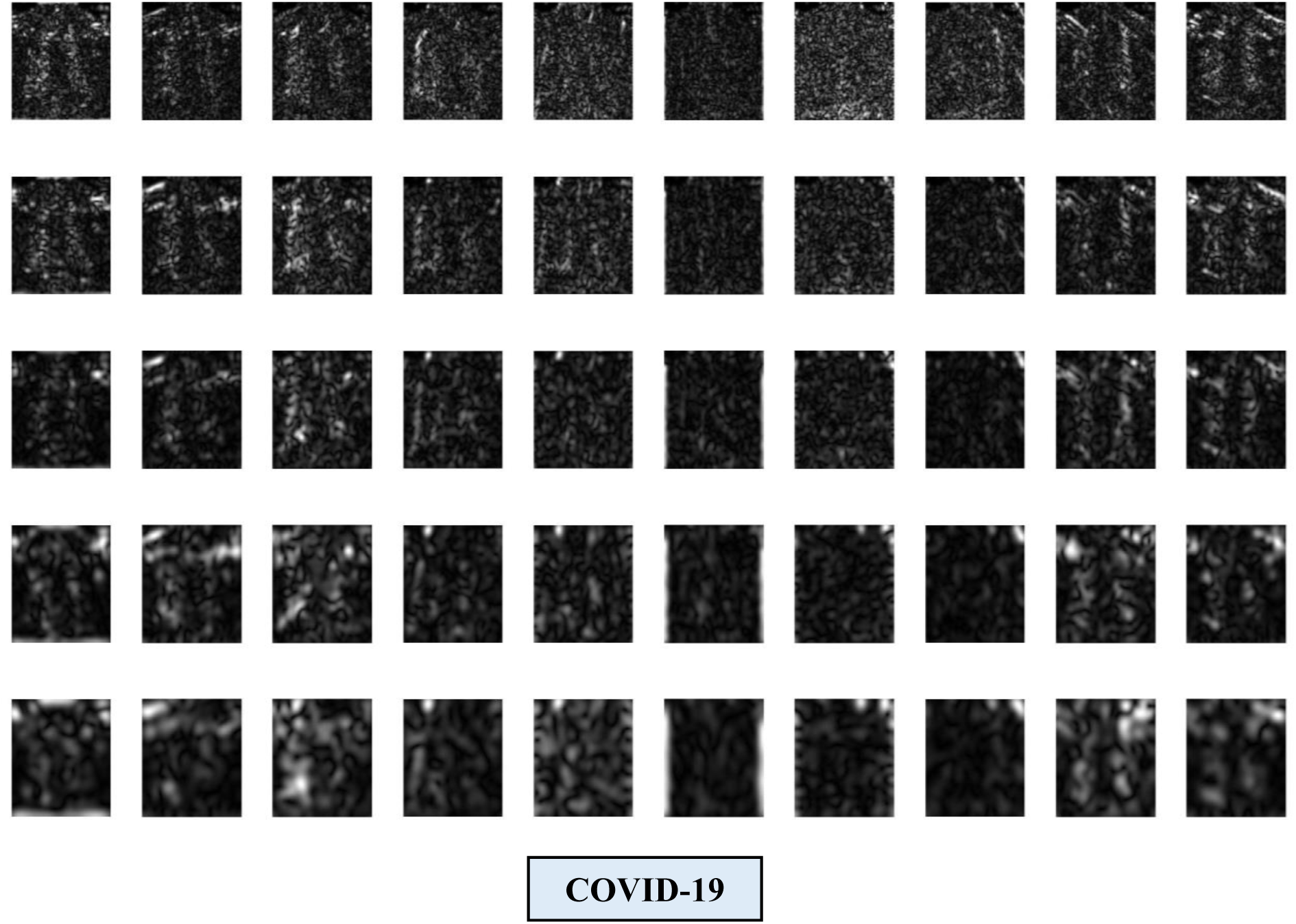

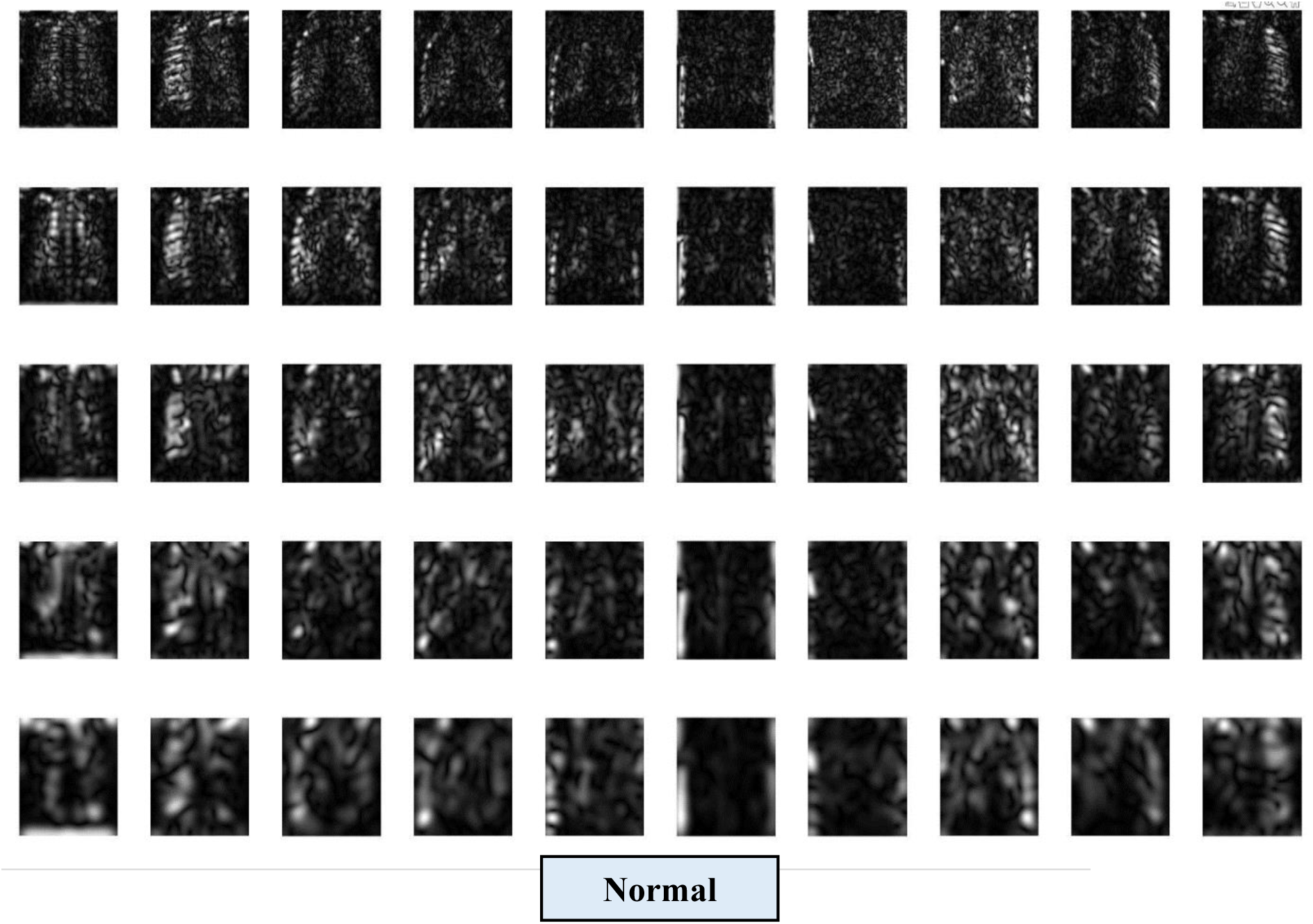

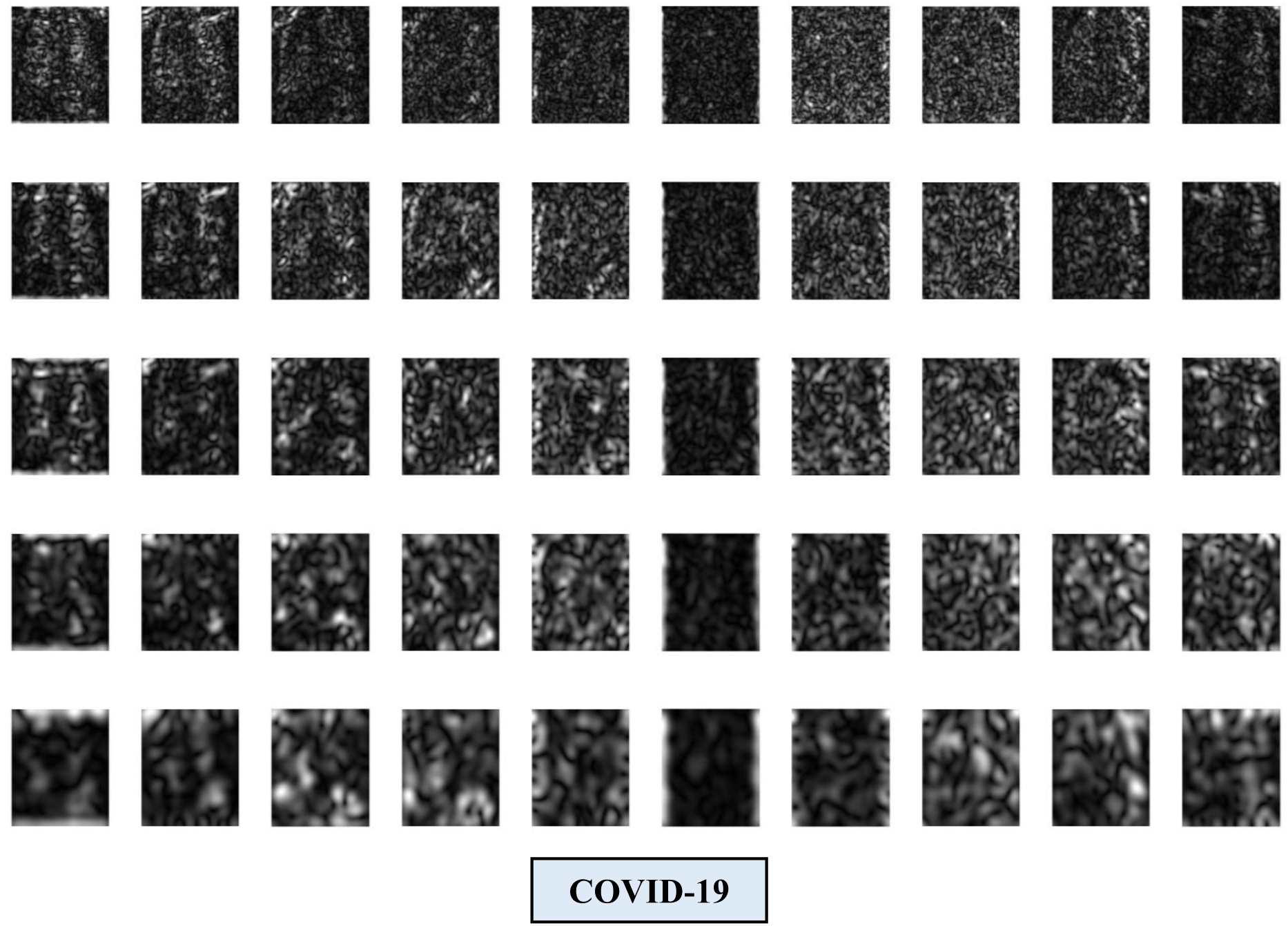

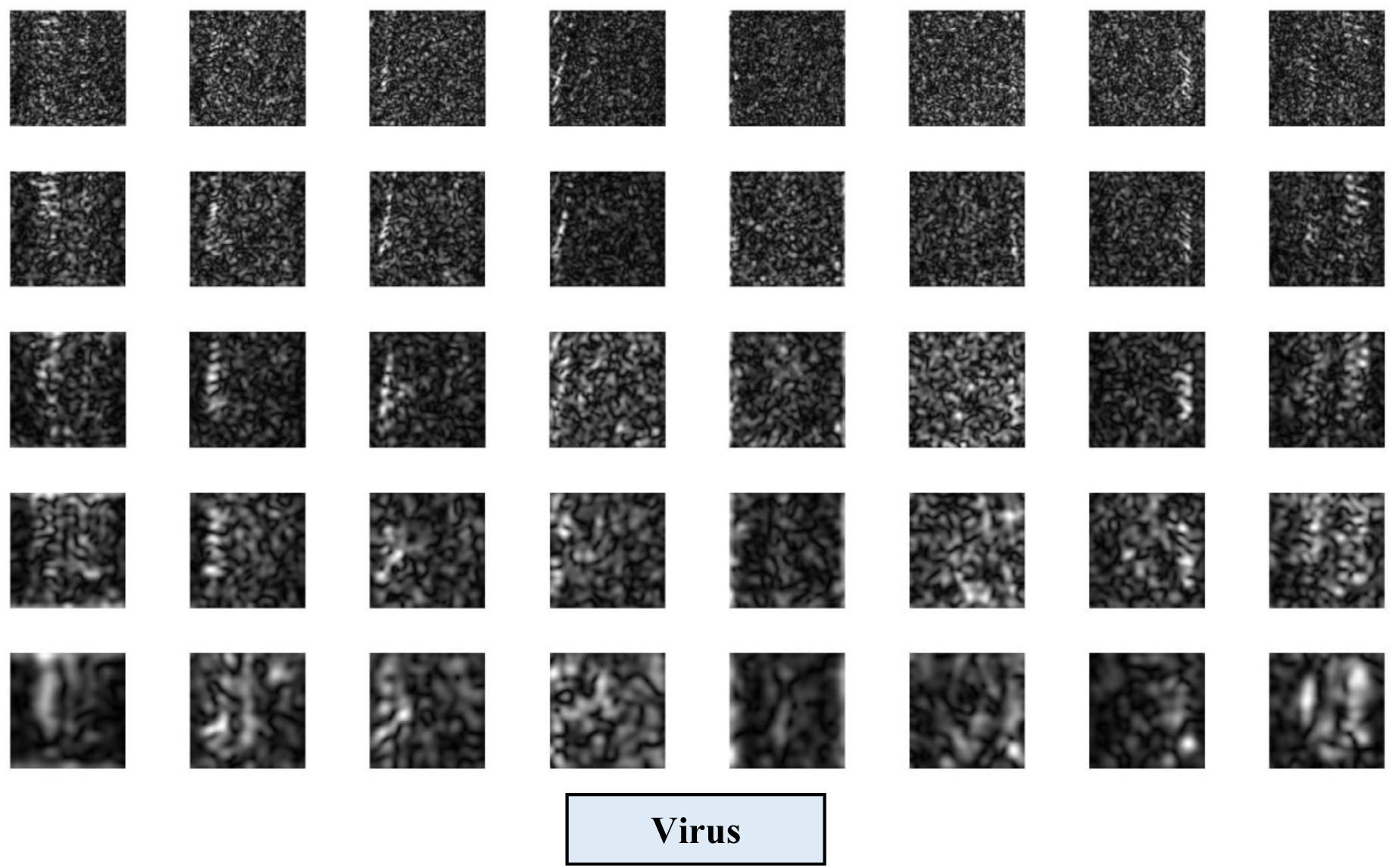

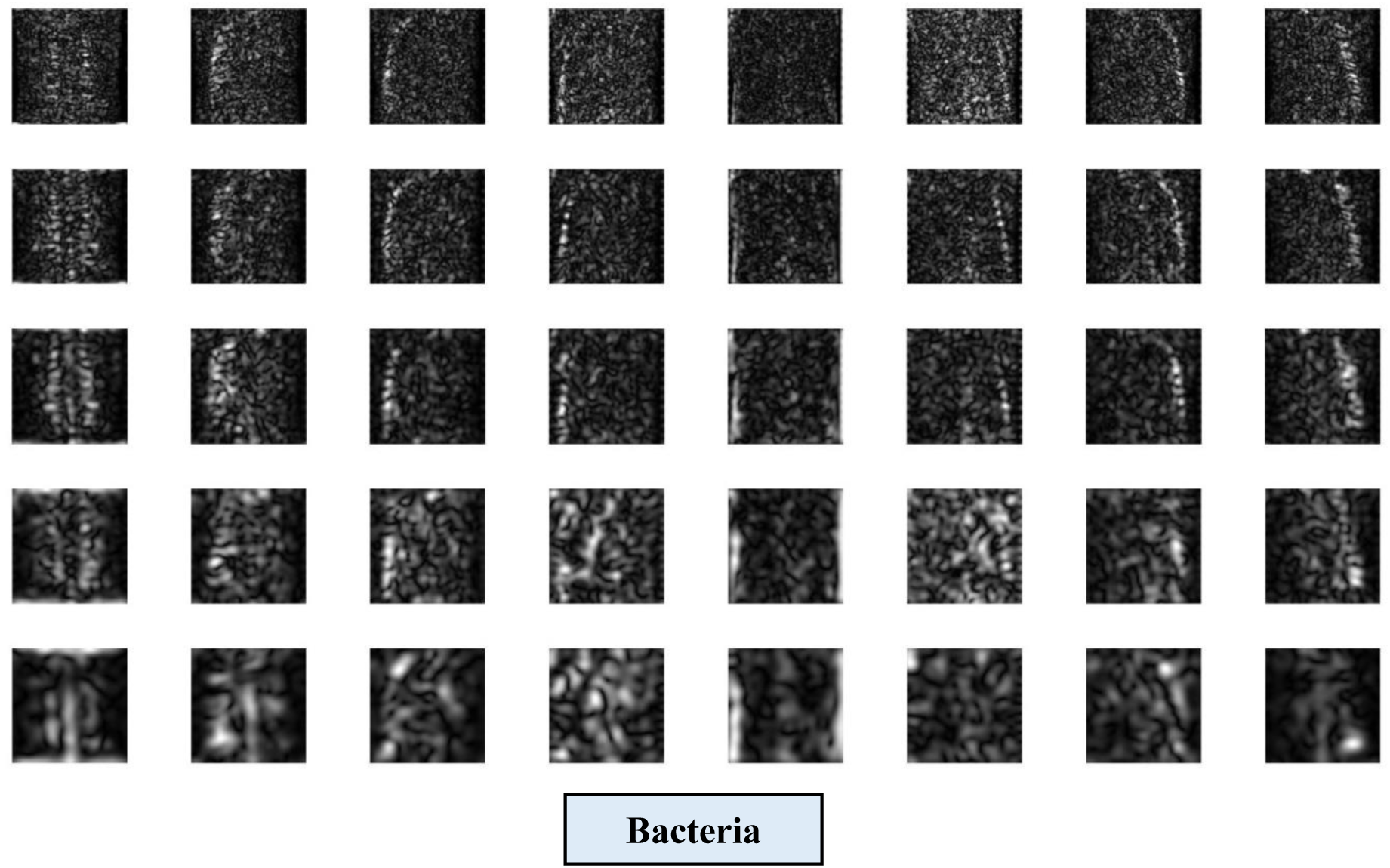

